# Temporal Immune Profiling in the CSF and Blood of Patients with Aneurysmal Subarachnoid Hemorrhage

**DOI:** 10.1101/2024.08.16.24312086

**Authors:** TA Ujas, KL Anderson, J Lutshumba, SN Hart, J Turchan-Cholewo, KW Hatton, AD Bachstetter, BS Nikolajczyk, AM Stowe

## Abstract

**Background:** Delayed cerebral ischemia (DCI) is a significant complication of aneurysmal subarachnoid hemorrhage (aSAH). This study profiled immune responses after aSAH and evaluated their association with DCI onset.

**Methods:** Twelve aSAH patients were enrolled. Leukocyte populations and cytokine levels were analyzed in cerebrospinal fluid (CSF) and peripheral blood (PB) on days 3, 5, 7, 10, and 14 post-aSAH. Peripheral blood mononuclear cells (PBMCs) were collected and their cytokine production quantified following stimulation.

**Results:** Mixed-effects models revealed distinct immune cell dynamics in CSF compared to blood. Natural killer T cell frequency increased over time in CSF only, while monocyte/macrophage numbers increased in both CSF and PBMCs. CD4+ HLA II+ T cells increased in circulation. Unstimulated PBMCs showed increased IL-1β, IL-6, and TNFα production, peaking at 7 days post-aSAH, coinciding with typical DCI onset. Ex vivo stimulation of PBMCs showed that only IL-6 significantly changed over time. In CSF, cytokines peaked 5 days post-injury, preceding immune cell profile alterations.

**Conclusions:** Our findings reveal a time-dependent immune response following aSAH, with distinct within-patient patterns in CSF and PB. The early CSF cytokine peak preceding immune cell changes suggests a potential mechanistic link and identifies the cytokine response as a promising therapeutic target. This cytokine surge may drive immune cell expansion and prime PBMCs for increased inflammatory activity, potentially contributing to DCI risk. Future studies should explore the importance and sources of specific cytokines in driving immune activation. These insights may inform the development of targeted immunomodulatory strategies for preventing or managing DCI in aSAH patients.

## Introduction

Subarachnoid hemorrhage (SAH), accounting for only 5% of all strokes [1], has a high pre-hospital mortality of 10-26% [2, 3], with an additional 35% mortality for those who reach the hospital [4]. A key complication following aneurysmal SAH (aSAH) is a vasospasm characterized by the constriction of intracranial vessels, leading to cerebral hypoperfusion and potential delayed cerebral ischemia (DCI) [5]. DCI involves cerebral and microvascular spasms, thrombosis, and cerebral autoregulation failure [6]. Up to 60-70% of individuals with aSAH experience vasospasm, with 40% developing DCI, typically occurring 4-14 days post-aSAH [7].

The pathogenesis of DCI includes RBC lysis, which releases toxic-free hemoglobin. Neutrophils and macrophages rapidly infiltrate the subarachnoid space, microvasculature, and brain parenchyma, phagocytosing RBCs and hemoglobin [8-12]. These cells degrade hemoglobin, the metabolites methemoglobin, heme, and hemin, which act as damage-associated molecular patterns (DAMPs). These DAMPs activate microglia via a TLR4-dependent pathway and endothelial cells through a TLR4-independent pathway, producing pro-inflammatory cytokines like TNFα, IL-1, and IL-6 [13-15]. Within 2-4 days post-aSAH, endothelin and oxygen-free radicals further promote inflammation and cerebral vasoconstriction. Hemoglobin directly damages neurons and reduces nitric oxide, impairing vasodilation and autoregulation and increasing DCI risk [16-18]. The ratio of neutrophils to lymphocytes at admission predicts DCI development [19].

While the role of neutrophils in DCI is increasingly understood, the contribution of lymphocytes remains largely unknown [20]. Recent observations have associated CD8^+^CD161^+^ T cells with vasospasm in aSAH [21], suggesting a more complex immune response than previously recognized. Despite these insights, longitudinal changes in immune response following aSAH and their relationship to DCI onset remain poorly characterized. Understanding these dynamics could provide opportunities for developing targeted immunotherapeutics to counter DCI.

This study aimed to profile longitudinal changes in immune responses following aSAH and evaluate their associations during typical DCI onset. We examined leukocyte populations and cytokine levels in CSF and peripheral blood at multiple time points post-aSAH. Additionally, we assessed cytokine production from stimulated peripheral blood mononuclear cells (PBMCs) to distinguish between aSAH-induced activation and intrinsic changes in immune function. Our findings highlight early cytokine production and the behavior of NK T cells and monocytes/macrophages in CSF compared to peripheral blood. Understanding innate and adaptive immune responses in different compartments post-aSAH provides a foundation for developing targeted therapies. This research aims to improve aSAH recovery management during critical weeks post-injury, potentially leading to better outcomes for individuals affected by this condition.

## Methods

### Data Availability

All data generated in this study are available in Table S3. Additional data are available following reasonable request.

All samples were collected from study participants with aSAH after approval by the University of Kentucky Institutional Review Board (IRB# 65452) and in accordance with the Declaration of Helsinki. All subjects and/or their authorized representatives signed written informed consent in accordance with Federal-wide Assurance on file with the Department of Health and Human Services (USA). All experiments were performed in accordance with relevant guidelines and regulations from 2020-2022.

### Subjects

Data were gathered from 12 study participants with aSAH aged 36-69 with aSAH. Age, sex, BMI, and patient demographic information were recorded, in addition to aneurysm location, Hunt and Hess (HH) score, and Fisher score. Vasospasm and DCI incidence were noted (see Table 1).

**Table 1.** Clinical and demographic characteristics of study participants.

| ID | Age | Gender | H&H | WFNS | Fisher | Aneurysm<br>Loc | EVD | Vaso | DCI |
| --- | --- | --- | --- | --- | --- | --- | --- | --- | --- |
| 1 | 50s | Male | 4 | 4 | 4 | Ant. | Yes | Yes | No |
| 2 | 30s | Female | 2 | 2 | 4 | Ant. | Yes | Yes | Yes |
| 3 | 60s | Female | 2 | 2 | 4 | Ant. | Yes | No | No |
| 4 | 40s | Female | 3 | 2 | 2 | Post. | Yes | Yes | No |
| 5 | 30s | Female | 2 | 1 | 3 | Ant. | No | Yes | No |
| 6 | 40s | Female | 2 | 2 | 2 | Ant. | No | No | No |
| 7 | 50s | Female | 2 | 1 | 4 | Ant. | Yes | Yes | Yes |
| 8 | 60s | Female | 2 | 1 | 3 | Post. | No | No | No |
| 9 | 60s | Female | 2 | 2 | 4 | Post. | Yes | Yes | Yes |
| 10 | 40s | Female | 3 | 2 | 4 | Post. | Yes | Yes | No |
| 11 | 30s | Female | 2 | 3 | 3 | Post. | Yes | Yes | No |
| 12 | 50s | Female | 2 | 2 | 3 | Post. | Yes | Yes | No |

The following abbreviations are used: ID (Study participant number), Age (Individual's age in years), Gender (Individual's gender), Ethnicity (Self-identified ethnic origin), H&H (Hunt & Hess Score, a grading system for subarachnoid hemorrhage severity), WFNS (World Federation of Neurosurgical Societies Score, assessing clinical condition in subarachnoid hemorrhage), Fisher (Fisher Score, predicting vasospasm risk in subarachnoid hemorrhage), Aneurysm Loc (Aneurysm Location, with "Post." indicating posterior and "Ant." indicating anterior locations), EVD (External Ventricular Drain presence), Vaso (Vasospasm presence), and DCI (Delayed Cerebral Ischemia, a complication post-subarachnoid hemorrhage). Additional note on day of DCI occurrence: Patient IDs 2 and 9 had a DCI on day 5 post-aSAH, and patient ID 7 had a DCI on day 7.

### Sampling

All CSF and peripheral blood were collected at five time points up to 14 days from an aSAH injury. CSF samples were acquired on days 3, 5, 7, and 10 from the study participants with an external ventricular drain (EVD) placed for clinical indications. The CSF was slowly drawn from the EVD under continuous intracranial pressure monitoring in the morning. Standard aseptic techniques were followed. When applicable, a peripheral blood sample was collected from the same person on days 3, 5, 7, 10, and 14 days post-aSAH. Samples were not collected after the person was discharged based on clinical indication. A total of 29 CSF samples were obtained from 8 study participants with an EVD. Additionally, 37 peripheral blood samples were obtained from the study participants. We had 6 participants with “matched” samples, i.e., CSF and peripheral blood samples from the same day. Not all study participants had an EVD placed; thus, no CSF was collected from these individuals.

### Leukocyte subsets in cerebrospinal fluid and peripheral blood

Analyses of leukocyte populations were conducted by flow cytometry. CSF and peripheral blood samples were processed in the laboratory [22], and cells were counted using a Nexcelom Cellometer with ViaStain (AOPI) dye. Samples were aliquoted for a general immunophenotyping panel. Cells were washed in 1X PBS and incubated in Ghost Dye 780 (Tonbo Biosciences) with PBS in the dark at 4ºC for 30 minutes. Cells were washed twice in FACS buffer (PBS, 1% Bovine serum albumin, 0.01% sodium azide). Afterward, cells were blocked using a human FcR blocking reagent (Mitenyi Biotec) for 5 min at room temperature. Antibodies were added without washing and incubated in the dark at 4ºC for 30 min. Cells were washed twice in FACS buffer and fixed in 1% paraformaldehyde plus 0.1% EDTA on ice for 30 minutes. Sample acquisition was conducted using a FACSymphony (BD Biosciences) flow cytometer and BD FACS DIVA software. All gating and event analyses were performed in FlowJo v10 (BD biosciences). All samples were stained with a general immunophenotyping panel (Live/Dead, CD45^+^, CD19, CD3, CD4, CD8b, CD161, CD14, CD138, CD66b, CD11b, CD11c, CXCR3). Additionally, blood samples were analyzed using a second more specific B and T cell panel (FACS Panel 2 markers: Live/Dead, CD45, CD19, CD3, CD4, CD8b, CD11b, CD11c, HLA-I, HLA DR DQ DP, IL-21R, CCR3, CD25, CD23, CD69, CCR7, CD154, CXCR5) (Tables S1, S2). All samples had doublet exclusion based on FSC and SSC. Those singlets were then gated with a live/dead marker. Finally, all CD45^+^ live cells were analyzed using uniform manifold approximation projection (UMAP). See supplement figure 1. For detailed step-by-step methods, please refer to “A Guide on Analyzing Flow Cytometry Data Using Clustering Methods and Nonlinear Dimensionality Reduction (tSNE or UMAP)” [23]. Non-linear dimensionality reduction and the FlowSOM clustering algorithm were used to visualize and interpret the flow cytometry data. FlowSOM runs an unsupervised clustering algorithm as a visualization aid to gain insight into subpopulations and their characteristics. FACS data was exported to an Excel file and analyzed for statistical significance. Note that the CSF had a unique granulocyte population, likely secondary to the immediate processing of CSF samples compared to the banking process and SepMate™ PBMC isolation protocol [24, 25].

**Figure 1:**
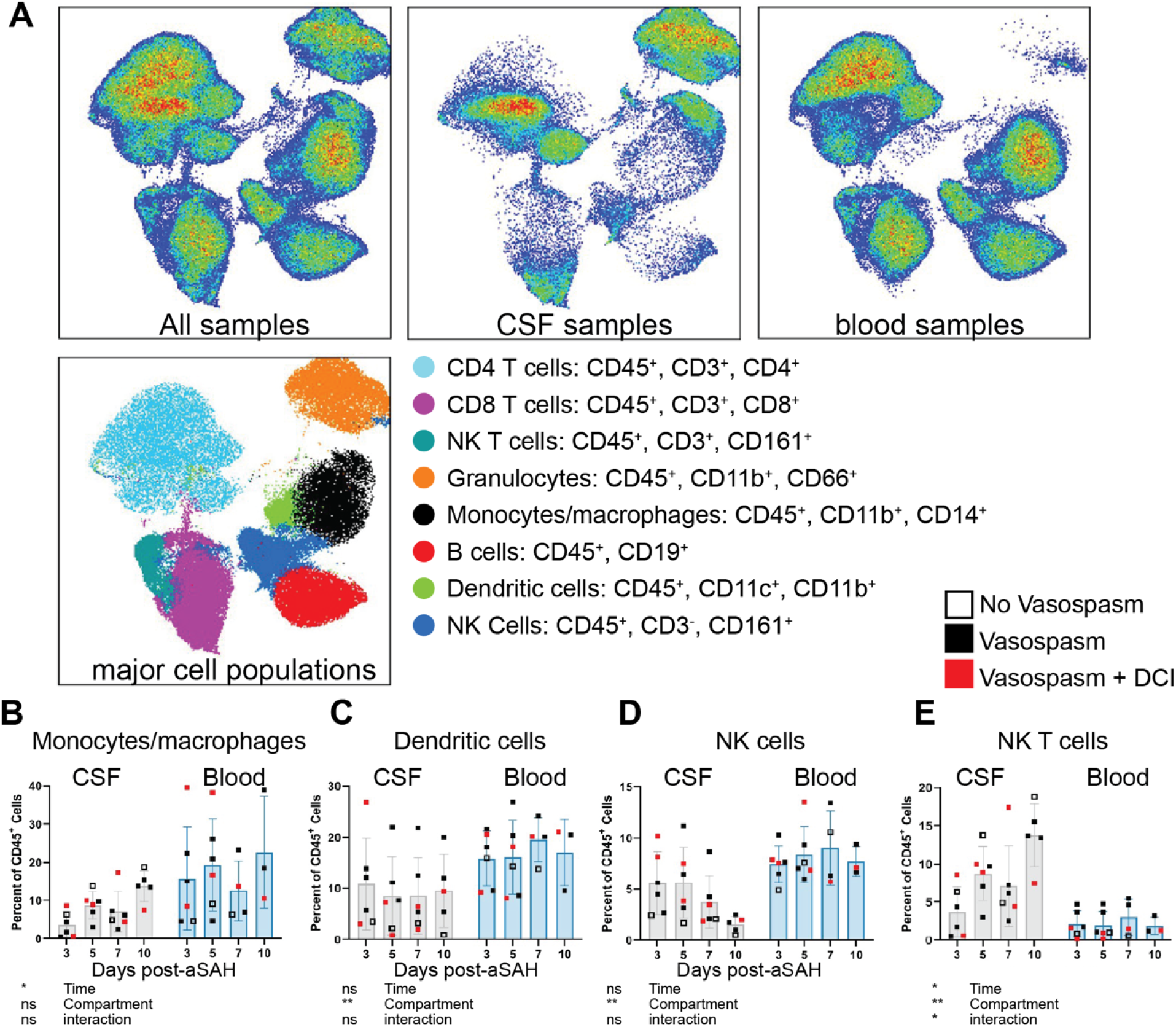
Central (CSF) versus peripheral (blood) immune cell populations show heterogeneity and temporal patterns post-aSAH. **(A)** UMAP plot of all samples, CSF, and peripheral blood samples showing the heterogeneity of immune cell populations. The analysis includes 6 study participants with matching CSF and peripheral blood samples. The main effects of time, fluid compartments, and their interaction were observed for **(B)** monocytes/macrophages, **(C)** dendritic cells, **(D)** NK cells, and **(E)** NK T cells. Visual comparison of study participants who developed vasospasm without (solid black) or with DCI (solid red) versus those who did not (open square) shows no clear pattern indicating the final diagnosis. Cell populations are shown as a percent of live CD45^+^. Bar graphs plot mean values; error bars indicate SD. The main effects are from a 2-way ANOVA (mixed-effects model REML) with Geisser-Greenhouse correction. *P<0.05; **P<0.01; ns = not significant.

### Cytokine production by stimulated PBMCs

The PBMCs from the study participants were collected as described above, with further methods described here [26, 27]. Each sample was resuspended in RPMI-1640 at 10^6^ cells/mL, seeded into 24-well plates (CellStar) at 250,000 cells/well, then either unstimulated or stimulated with E. Coli O111:B4 LPS (25 ng/mL, Millipore Sigma) to primarily target myeloid cells, or stimulated human αCD3/αCD28 Dynabeads (1 bead/cell, Gibco) to primarily target T cells, for 20 and 40hrs, respectively. Cytokines were quantified using a 25-plex Th17 magnetic bead kit (Millipore Sigma) and a Bio-Rad FLEXMAP 3D with Luminex xPONENT 4.2 and Bio-plex Manager (Bio-Rad) software with 1:10 dilutions as appropriate. Values below the detection level were replaced with a value of 1/10 of the minimum standard curve value (specific to each cytokine) for analysis by approaches requiring minimum values of >0.

### Cytokine protein assays

We used CSF samples from 8 study participants to quantify cytokines MSD S-plex Proinflammatory panel (K15396S), MULTI-SPOT for 9 cytokines: IFN-γ, IL-1β, IL-2, IL-4, IL-6, IL-10, IL-12p70, IL-17A, TNF-α. All samples were run 1:1 with blocking solution and sample volume, according to the MSD S-plex protocol.

### Statistical analysis

Analyses were performed using GraphPad PRISM or JMP Pro Version 17.0.0. For Figure 1, a two-way ANOVA or Mixed-effects model (REML) was used to compare cell populations in CSF and PBMCs. Due to differing time points (14 days for PBMCs, 10 days for CSF), only a main effects-only model without Geisser-Greenhouse correction could be fitted. Figure 2 utilized a one-way Repeated measures ANOVA (REML). For Figure 3, a one-way repeated measures ANOVA (mixed-effects model REML) was applied to Log2 transformed data to measure the main effects. Student’s t-tests on Log2 transformed data compared day 3 vs. day 5, day 3 vs. day 7, and day 3 vs. day 10. Figure 4 analysis employed a one-way repeated measures ANOVA (mixed-effects model REML) on Z-score data, with Student’s t-tests comparing day 3 vs. day 5, day 3 vs. day 7, and day 3 vs. day 10. Z-scores were calculated from cytokine pg/ml values. Significance levels: *P<0.05, **P<0.01.

**Figure 2:**
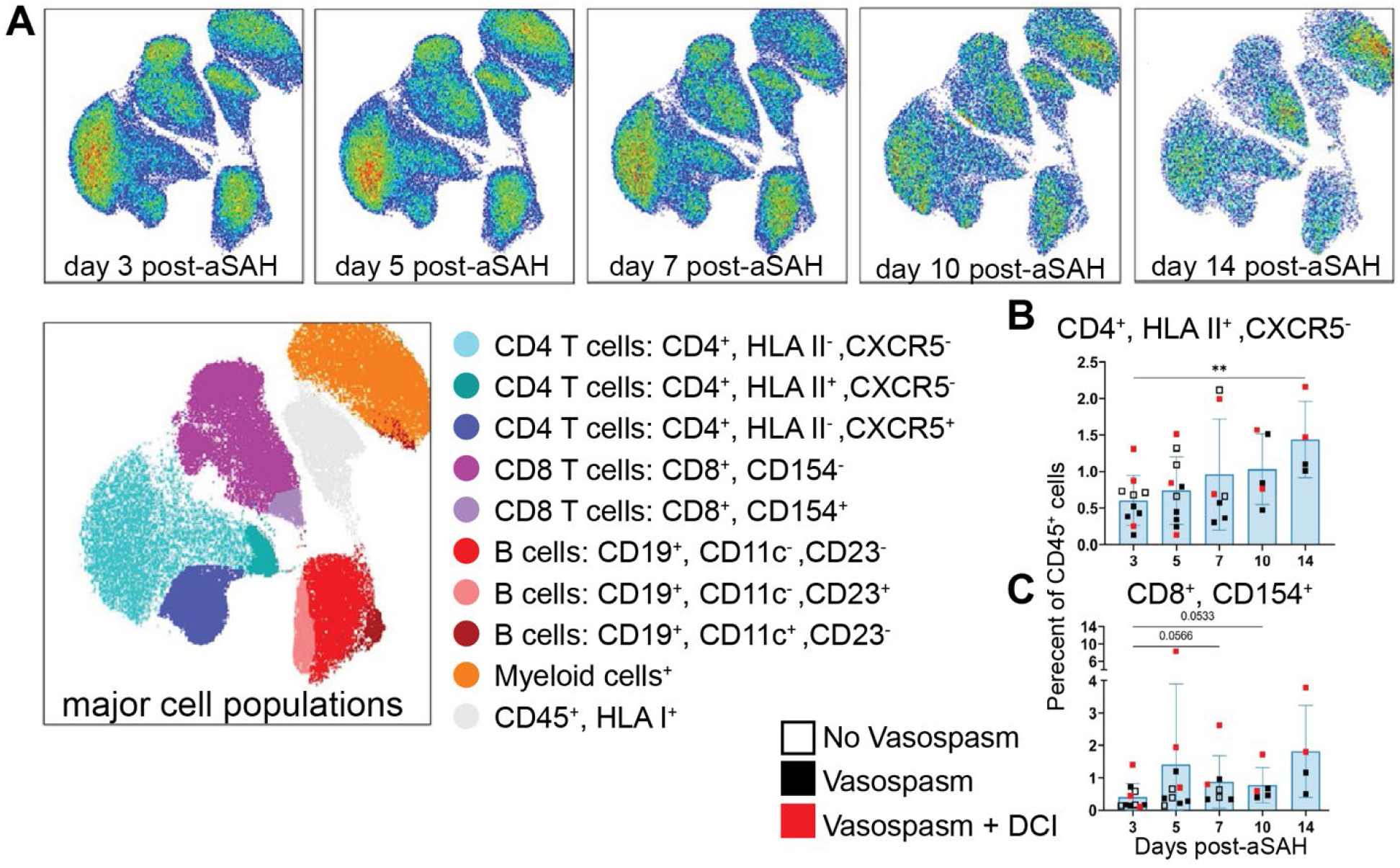
CD4 and CD8 T cell subpopulations increase in the blood over time after aSAH. **(A)** Flow cytometry UMAP plot of all samples. **(B)** A unique population of HLA class II^+^CD4^+^ T cells demonstrated an increase over time in peripheral blood. These cells continued to increase from days 5-14 post-aSAH, reaching the highest percentage on day 14 (vs. day 3, p=0.0089). **(C)** CD154^+^ CD8^+^ T cells trended higher from day 3 to day 7 (p=0.0566) and day 10 (p=0.0533) post-aSAH. Visual comparison of individuals who developed vasospasm without (solid black) or with DCI (solid red) versus those who did not (open square) shows no clear pattern indicating the final diagnosis. Cell populations are shown as a percentage of live CD45^+^ cells. Bar graphs plot mean values; error bars indicate SD. The main effects are from a one-way ANOVA (mixed-effects model REML) with Geisser-Greenhouse correction. Dunnett’s multiple comparisons test was conducted by comparing the means of each day to the mean from day 3. **P<0.01. Data is from 10 patients.

**Figure 3:**
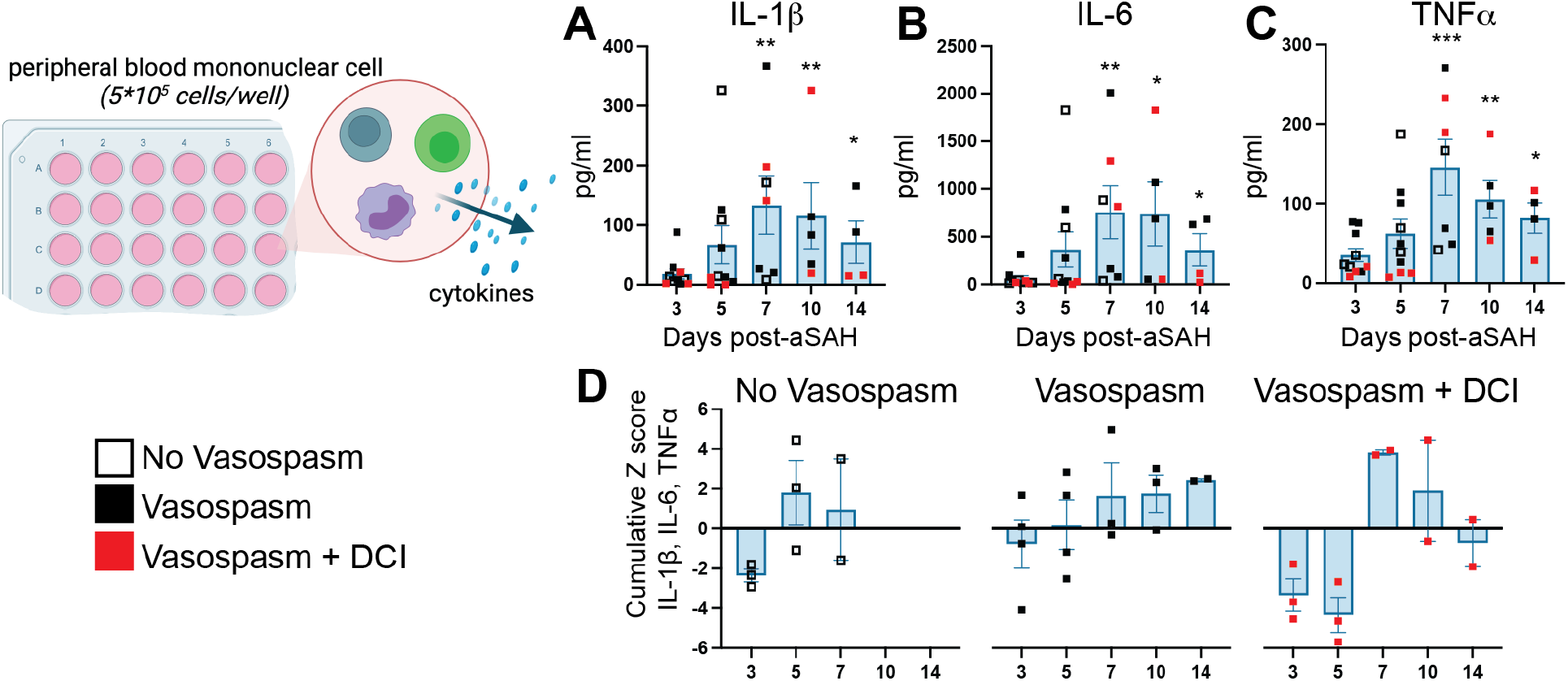
Basal cytokine production by PBMCs increases post-aSAH. Supernatants from unstimulated PBMCs incubated for 40 hours were measured 3-14 days post-aSAH. **(A)** IL-1β peaked around day 7 (p=0.0220). **(B)** IL-6 was significantly higher at day 7 (p=0.0236). **(C)** TNFα showed significant increases on days 7 (p=0.0149) and 10 (p=0.0452) compared to day 3. **(D)** A pro-inflammatory cytokine cumulative z-score of IL-1β, IL-6, and TNFα, stratified by vasospasm and DCI status, revealed a unique profile of proinflammatory changes. The Vasospasm+DCI group failed to activate a proinflammatory response to the injury at days 3-5 but showed a reversal at day 7. Bar graphs plot mean values; error bars indicate SD. The main effects were analyzed using a one-way repeated measures ANOVA (mixed-effects model REML). Student’s t-tests were used to compare day 3 vs. day 5, day 3 vs. day 7, and day 3 vs. day 10. *P<0.05, **P<0.01, ***P<0.001. Data is from 10 patients. See also Table S3 for all analytes measured.

**Figure 4:**
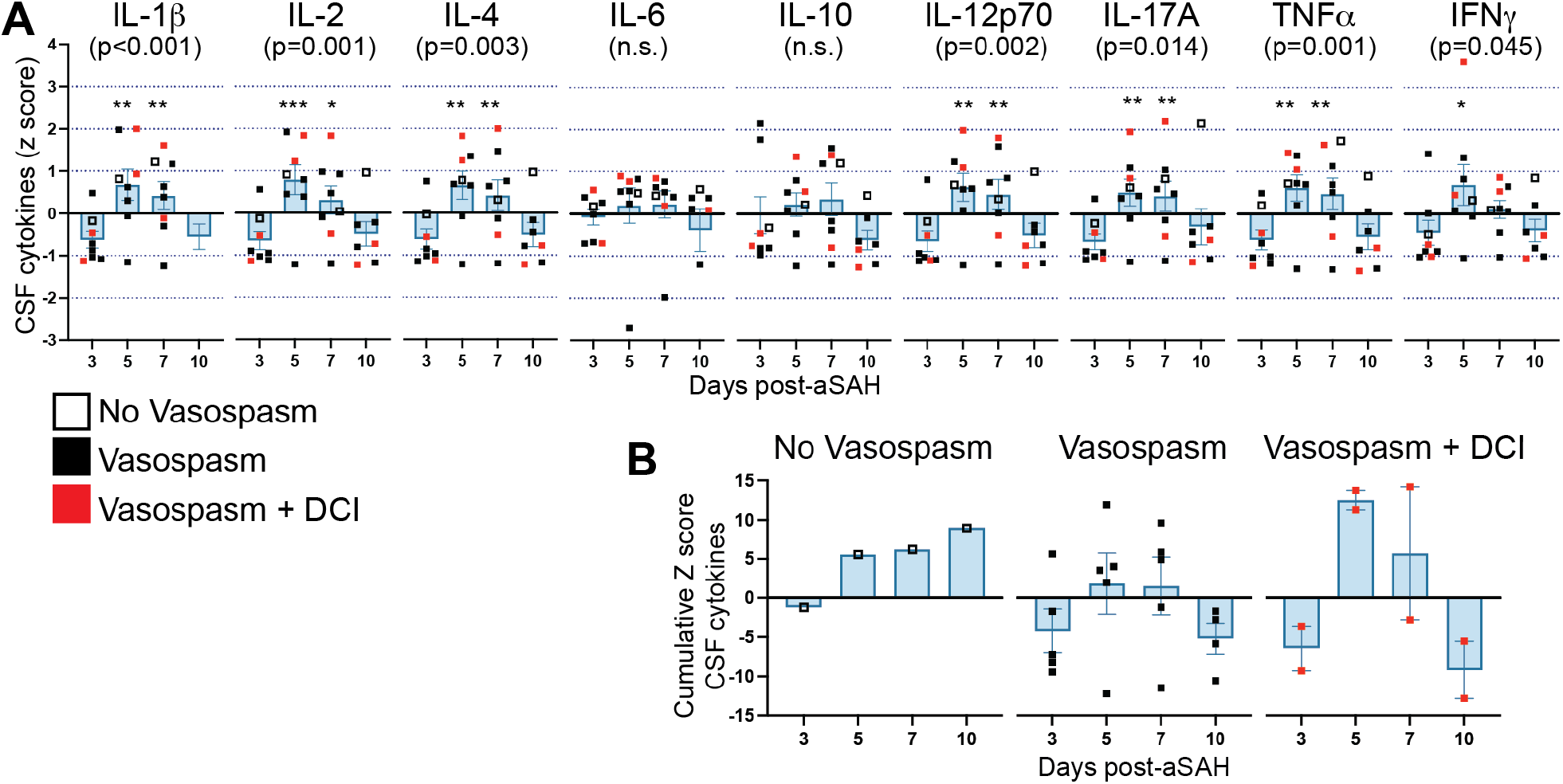
Temporal dynamics of CSF cytokine levels following aSAH. **(A)** Cytokine levels were measured in CSF at days 3, 5, 7, and 10 post-aSAH. IL-1β, IL-2, IL-4, IL-12p70, IL-17A, TNFα, and IFNγ showed a significant main effect of time, with peak levels occurring at day 5 post-aSAH. **(B)** Cumulative z-score of measured cytokines over time, stratified by vasospasm and DCI status. A main effect of time was observed (F=5.74, p=0.005), with days 5 and 7 significantly elevated compared to day 3 (p=0.004 and p=0.017, respectively). The vasospasm+DCI group showed a distinct pattern with low cytokine levels at day 3, followed by a marked elevation at day 5. Bar graphs represent mean values; error bars indicate SD. The main effects were analyzed using a one-way repeated measures ANOVA (mixed-effects model REML). Student’s t-tests were used to compare day 3 vs. day 5, day 3 vs. day 7, and day 3 vs. day 10. *P<0.05, **P<0.01. n =8. Z-scores were calculated from the pg/ml values of cytokines. Raw pg/ml values and z-scores for all cytokines are provided in Table S3

## Results

The clinical and demographic characteristics of the twelve enrolled study participants are shown in **Table 1**. The study participants had a median age of 51 and a median BMI of 31.85. Most study participants (9) presented with a Hunt and Hess (HH) score of 2. Half of the study participants had a Fisher grade 4 aSAH (n=6), with 2 study participants having a grade of 2 and 4 with a grade of 3. Nine study participants (75%) developed vasospasm and 3 (25%) developed DCI. Additionally, 3 study participants (25%) presented with additional aneurysms.

### Distinct immune cell clusters can be found in the CSF compared to peripheral blood following aSAH

Clustering analysis reveals distinct immune cell profiles in CSF and peripheral blood (**Figure 1**). NK T cells, a subset of CD4+ T cells, CD8+ T cells, B cells, and monocyte/macrophages are more abundant in peripheral blood. In contrast, granulocytes and a subset of CD4+ and CD8+ cells are more abundant in CSF (**Figure S2**). Comparison of the temporal patterns of immune cell populations in CSF and peripheral blood after aSAH in 6 patients showed that the percentage of monocytes and macrophages increased with time post-aSAH (p=0.0373, **Figure 1B**), with a more pronounced increase in CSF. DCs showed no temporal pattern but were more frequent in peripheral blood than CSF (p=0.0032, **Figure 1C**). NK cells were found in greater frequency in peripheral blood (p=0.0016) but were not affected by time in either compartment (**Figure 1D**). Interestingly, NK T cells showed an increased frequency over time only in CSF (time x compartment interaction: p=0.0263, **Figure 1E**), with higher levels in CSF compared to peripheral blood (p=0.0095). Global CD19^+^ B cells (p=0.0041, **Figure S2**) and CD8^+^ T cells (p=0.0458, **Figure S2**) were found at much lower levels in CSF than peripheral blood without temporal changes. No changes were observed between compartments or over time for CD4^+^ T cells (**Figure S2**). The changes in immune cell numbers around the 5-10 day timepoint correspond temporally to when DCI develops post-aSAH [7] though comparison between patients with and without vasospasm, or the 3 that developed DCI, do not show a clear pattern for driving any of the temporal changes (**Fig 1B-E**).

While we did not see temporal changes in global B cell or CD4 or CD8 T cells, the heterogeneity seen in our UMAP analysis (**Figure 1A**) within these cell populations suggested that exploring these populations, particularly in the peripheral blood, in more depth could identify unique B and T cell populations missed in our primary analysis. Using a more focused lymphocyte panel (**Table S2**), we identified several additional clusters of B and T cells (**Figure 2A**). Of these clusters, we found that CD4^+^ HLA class II^+^ T cells (p=0.0089, **Figure 2B**) and CD8^+^CD154^+^ T cells (p=0.0566, **Figure 2C**) were increased in the peripheral blood over time post-aSAH. The other populations, including CD19^+^CD11c^+^ B cells, showed no main effect of time after aSAH (**Figure S3**).

### Increased basal cytokine production by PBMCs post-aSAH without changes in stimulated response

Next, we completed immunophenotyping of PBMCs to understand if differences in function and number were seen after aSAH. We found a main effect of time post-aSAH for TNFα (**Figure 3A**, p=0.004), IL-1β (**Figure 3B**, p=0.014), IL-6 (**Figure 3C**, p=0.014), and IL-10 (**Figure S4**, p=0.019) released into the media after 40h of culture without stimulation. To better visualize the overall pro-inflammatory response, we calculated a cumulative z-score of TNFα, IL-1β, and IL-6 (**Figure 3D**). This analysis, when stratified by vasospasm and DCI status, revealed a unique profile of proinflammatory changes, with the Vasospasm+DCI group showing a delayed activation of the proinflammatory response until day 7 post-aSAH. However, it is important to note that the small sample size, particularly when stratified into subgroups, precludes the use of statistical analyses. Following LPS stimulation, no effect of time after aSAH was seen for any of the cytokines though, as expected, levels were 10-100-fold higher with *E. coli* LPS stimulation than in the unstimulated cells (**Figure S4**). Following T cell stimulation with CD3/CD28, IL-6 was the only cytokine that exhibited temporal change (**Figure S4**), with a main effect of time (p=0.048, **Figure S4**) that peaked around 10d post-aSAH, which agreed with data from the unstimulated cells. The ∼3-fold increase in IL-6 following stimulation likely primarily reflects the response from the unstimulated cells (**Figure S4**).

### A distinct cytokine profile emerges in CSF 5 days post-aSAH

To evaluate drivers of immune cell changes, we measured cytokine levels in the CSF at days 3, 5, 7, and 10 post-aSAH (**Figure 4A**). We found a significant main effect of time after aSAH for IL-1β, IL-2, IL-4, IL-12p70, IL-17A, TNFα, and IFNγ. A conserved pattern emerged across cytokines, with peak activation occurring 5 days post-aSAH. The cytokines were highly correlated, with an average Pearson’s R of 0.79 and a minimum of 0.55. Reducing the dimensionality of the data using a cumulative z-score of the cytokines measured in **Figure 4A**, we found a main effect of time after injury (F=5.74, p=0.005), with days 5 and 7 post-aSAH significantly elevated over day 3 (p=0.004, p=0.017, respectively).

In an exploratory analysis, we examined patterns in CSF cytokines associated with vasospasm and DCI. Similar to PBMC findings, we observed a greater fluctuation in study participants with vasospasm+DCI, which was not apparent in other groups (**Fig 4B**). On day 3, cytokine levels were low in the CSF of the vasospasm+DCI group, but they showed a marked elevation on day 5 that exceeded other cytokine levels observed. The timing is noteworthy as changes in CSF cytokine levels occur at least 2 days before the PBMC responses, supporting the hypothesis that CSF cytokines may drive systemic immune changes following aSAH.

## Discussion

DCI remains a key complication after aSAH and a major cause of morbidity and mortality [28]. The intricate immune response following aneurysmal rupture may provide significant opportunities for immunotherapeutics to counter DCI. Thus, the goal of the present study was to longitudinally characterize the immune response following aSAH, aiming to identify CSF-based populations associated with DCI. Temporally, we found increases in immune cell populations and cytokines in the CSF that correspond to the typical onset of DCI around 4-14 days post-aSAH [7], suggesting that elevated inflammation could contribute to vasospasm and DCI. We also observed basal activation of PBMCs with increased IL-1β, IL-6, and TNFα expression. A unique temporal pattern was seen for the vasospasm+DCI group compared to other groups, notably showing a potential failure of early basal activation by aSAH during the first 5 days after injury. However, the small sample size limits statistical confirmation, particularly regarding the potential failure of basal activation in the vasospasm+DCI group during the early post-injury period.

For innate populations, our current study found that monocytes and macrophages continued to increase in the CSF over post-aSAH days 3-10, confirming prior work [29]. The role of infiltrating monocytes and macrophages in SAH is poorly understood due to the difficulty distinguishing these innate cells from microglia and perivascular macrophages in the CNS [11], though our results clearly suggest a continued recruitment and/or proliferation of these innate immune cells in the CSF following SAH. Previous studies report that NK cells in CSF initially increase, then decline in individuals with aSAH with vasospasm [30], also aligning with our observation of lower NK cell numbers at day 10. Lower acute NK cell numbers were linked to vasospasm [30], but our small sample showed the opposite trend. In contrast, we found NK T cells increased over time in CSF but not in blood. NK T cells are known to infiltrate brain tissue after cerebral ischemia, producing IFNγ, IL-17, IL-4, and IL-10 [31, 32], and we previously found them increased in intracranial blood within 24 hours of ischemic stroke onset [22]. Our study shows NK T cells continue infiltrating or proliferating in the CNS days after aSAH concomitant with monocytes/macrophages, though the role of NK and NK T cells in aSAH remains poorly characterized, warranting further investigation.

Unlike in ischemic stroke [22], data on lymphocytes in aSAH is scarce [11, 20]. Relevant experimental models show CD3^+^ T cells infiltrating the injured hemisphere as early as day 1 and days 4-5 post-injury [33-35], with CD3^+^CD4^+^ T cells being the predominant brain-infiltrating lymphocyte subpopulation [35, 36]. Our study found that pan CD4^+^ T cell levels were similar between CSF and blood, with no change over time. However, we observed an increase in CD4^+^HLAII^+^CXCR5^-^ cells in the blood after aSAH, peaking 14 days post-injury. Nonprofessional antigen-presenting cells (APCs), such as T cells, only acquire MHC class II expression under unique circumstances [37]. While these cells may contribute to T lymphocyte activation via cytokine production and antigen presentation, their specific function in aSAH remains undefined. Interestingly, one study found that these antigen-presenting T cells predominantly differentiate into regulatory T cells [38], which can positively impact recovery after ischemic stroke [39].

We observed no temporal changes in pan CD8^+^ T cells but found higher blood levels than CSF, also confirming prior studies [29, 40]. A subtype of CD8^+^CD154^+^ T cells increased over time post-aSAH, supporting previous work showing heterogeneity in the CD8 T cell response to aSAH. A study of 13 individuals with aSAH found that CD8^+^CD161^+^ T cells (cytotoxic Tc17 cells that secrete IL-17) increased significantly in the CSF of individuals with vasospasm [30]. While we did not characterize this specific subset, we analyzed CSF IL-17 levels and found significant elevations at post-aSAH days 5 and 7. Given that IL-17’s main function is to recruit monocytes to inflammation sites by increasing local chemokine production [41], it is noteworthy that we observed continued monocyte/macrophage increases in CSF following IL-17 upregulation. B cells are the remaining pan-lymphocyte population and can be either neuroprotective in acute stroke phases or contribute to cognitive impairment chronically [42, 43]. In experimental models, B cells support neurogenesis and functional recovery [44] and infiltrate the brain post-injury [36]. In aSAH patients, oligoclonal immunoglobulin bands in serum, and CSF suggests polyclonal B cell activation [45]. We observed higher B cell levels in blood than CSF but no significant temporal changes in B cell populations, which contrasts with some studies and underscores the need for further research to clarify B cells’ contributions in aSAH progression [29, 46].

In the CSF, cytokines (TNFα, IL-1β, IL-2, IL-4, IL-12, and IL-17) peaked 5 days after injury, preceding alterations in immune cell profiles. This sequence suggests that the cytokine response may drive the expansion of immune cell populations in both CSF and peripheral blood, potentially priming PBMCs to become more inflammatory and increase cytokine production. In response, PBMCs produced proinflammatory cytokines (IL-1β, IL-6, and TNFα) without additional stimulation, confirming that aSAH activated PBMCs, with peak production occurring during the time window when vasospasm and DCI are most likely to occur. Unexpectedly, in study participants with post-aSAH vasospasm+DCI, there was a marked change in PBMC activation from day 5 to day 7. While the sample size is too small for definitive conclusions, this observation supports a potential involvement of cytokines in the development of vasospasm and DCI. Despite the basal activation of PBMCs after aSAH, their maximum activation after a strong immunological challenge was largely unaffected, suggesting that post-injury immunosuppression was not occurring within the time window of the current study. Finally, our findings align with others who found greater T cell-derived IL-1β in the CSF after SAH [47]. In relevant experimental models, blocking IL-1β mitigates early brain injury and BBB damage [48, 49] which may be an outstanding future target for immunotherapeutic development.

Our study has several limitations. First, the sample size (n=12) was too small to statistically test the involvement of CSF and blood immune changes in the development of vasospasm and DCI. Second, given the nature of the study and the standard of care for the participants, missing data for some participants was unavoidable. Demographically, our study was not balanced for gender, as only one male agreed to participate out of the 12 subjects. Finally, the study was based on a single site, which further limits the generalizability of our findings.

In summary, a diverse range of cells produce inflammatory cytokines that likely contribute to inflammation in aSAH. The compromised blood-brain barrier and presence of blood at the hemorrhage site present unique challenges, triggering a dynamic inflammatory response mediated by innate and adaptive immune cells. Our study highlights unique changes in monocytes/macrophages and NK T cells, which warrant further exploration, as well as a need for deeper profiling of B and T cells to reveal subpopulations contributing to the disease. The involvement of central (CSF) cytokines in modulating systemic (blood) immune function also merits further investigation. By documenting the immune system’s role in aSAH, we lay the foundation for future studies that can mechanistically explore ways to improve patient outcomes following aSAH through a large time window for administration of immunotherapies.

## Supporting information

STROBE checklist

## ARTICLE INFORMATION

**Supplemental Material**

**Figure S1-S4**

**Tables S1-S3**

## Acknowledgments

Our sincere thanks go out to all the research participants and their families. Some illustrations were created using BioRender.com. The research was supported by the University of Kentucky Neuroscience Research Priority Area. As well as National Institutes of Health grants: T32AG057461 (JL), R01NS103785 (ADB), R56AG069685 (BSN), R56AG074613 (AMS), R01NS122119 (AMS) and AHA 19EIA34760279 (AMS). The project described was supported by the NIH National Center for Advancing Translational Sciences through grant number UL1TR001998. The content is solely the responsibility of the authors and does not necessarily represent the official views of the NIH.

**Footnote**

## Nonstandard Abbreviations and Acronyms

SAH: Subarachnoid hemorrhage
aSAH: Aneurysmal Subarachnoid Hemorrhage
CSF: cerebrospinal fluid
DCI: Delayed cerebral ischemia
PB: peripheral blood
PBMCs: Peripheral blood mononuclear cells
RBC: Red blood cell
EVD: external ventricular drain
HH score/ H&H/ HH: Hunt and Hess score
Fisher: Fisher grading scale for subarachnoid Hemorrhage
WFNS: The World Federation of Neurological Surgeons grading scale for subarachnoid hemorrhage
Aneurysm Loc: Aneurysm Location
Ant.: Anterior
Post.: Posterior
Vaso: Vasospasm presence
FACS: fluorescence-activated cell sorting
UMAP: Uniform manifold approximation and projection
LPS: Lipopolysaccharides
BBB: blood–brain barrier
APCs: antigen-presenting cells

