## Supplemental figures for "Temporal Immune Profiling in the CSF and Blood of Patients with Aneurysmal Subarachnoid Hemorrhage"

1-Department of Neuroscience, University of Kentucky, Lexington, USA
2-Department of Neurology, University of Kentucky, Lexington, USA
3-Department of Anesthesiology, University of Kentucky, Lexington, USA
4-Univ Kentucky, Spinal Cord & Brain Injury Res Ctr, 741 S Limestone St, Lexington, KY 40536 USA
5-Sanders-Brown Center on Aging, University of Kentucky, Lexington,
USA
6-Department of Pharmacology and Nutritional Sciences, University of Kentucky Lexington, Kentucky,
USA
7-Barnstable Brown Diabetes Center, University of Kentucky Lexington, Kentucky, USA

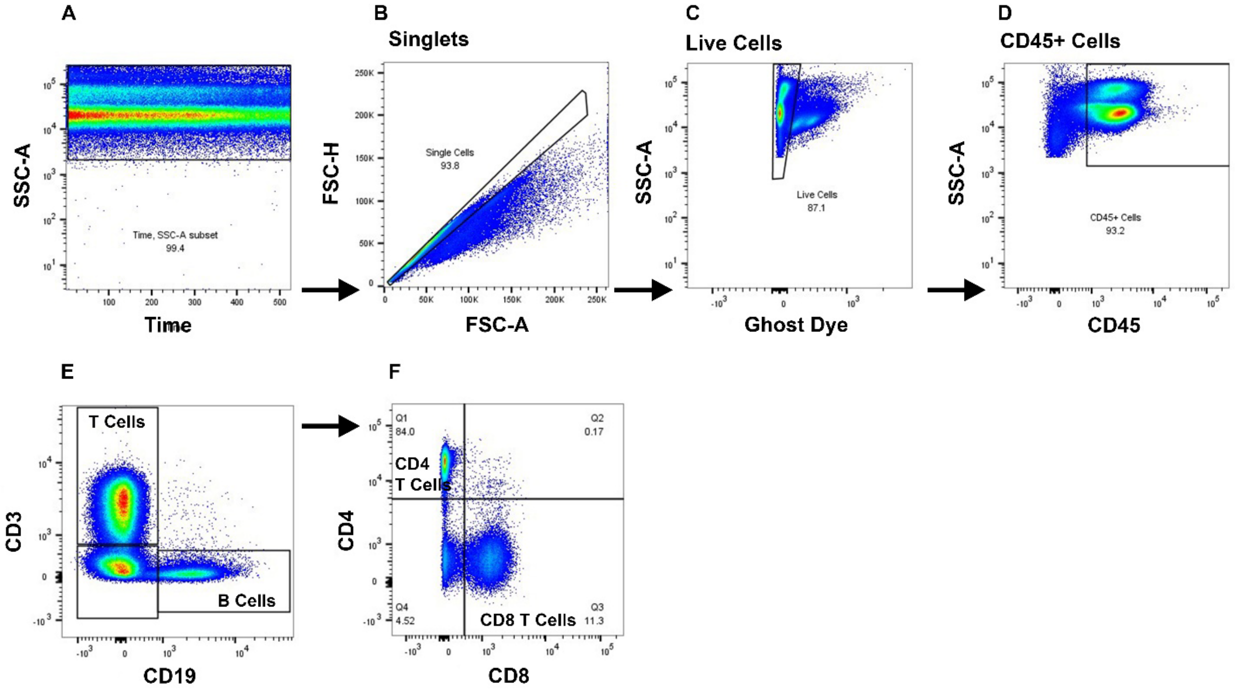


**Supplemental Figure 1: Flow cytometry gating strategy.** Samples were gated as shown in Supplemental Figure 1A-D, with numeric tags assigned for later unblinding. Live CD45+ cells were concatenated for high-dimensional analysis. UMAP was used to reduce high-dimensional data to two dimensions, followed by FlowSOM for unbiased clustering on the dimensionality reduction map. This approach yielded automatic cluster gates revealing unique populations, as shown in Figure 1. Cell populations from FlowSOM-driven meta clustering were identified based on markers listed in Figure 1's legend. Supplemental Figure 1E-F display traditional gating of CD4 and CD8 T cells for verification of FlowSOM meta clusters.

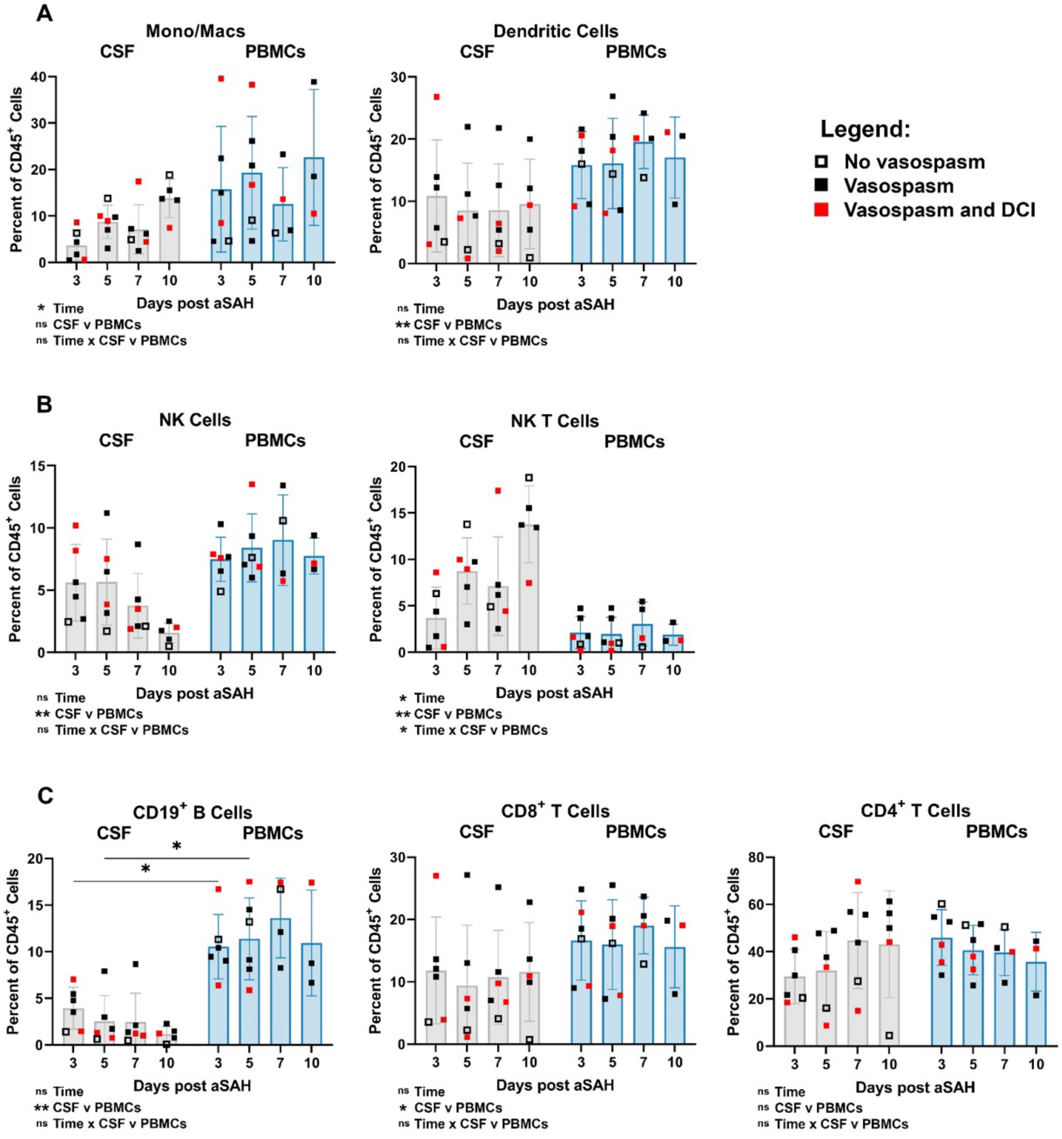


**Supplemental Figure 2: Comparison of immune cell populations in CSF and PBMCs from aSAH patients.** A) Innate immune cells. B) NK cells and NK T cells. C) Lymphocytes. Bar graphs represent mean values; error bars indicate SD. Cell populations are shown as a percentage of live CD45+ cells. Hollow squares indicate patients without vasospasm, filled squares indicate patients with vasospasm, and red squares indicate patients with vasospasm and delayed cerebral ischemia (DCI). A 2-way ANOVA (mixed-effects model REML) with Geisser-Greenhouse correction was performed, followed by Tukey's multiple comparisons test for each comparison. Fixed effects (type III) are listed under each graph. Multiple comparisons were only conducted for corresponding days between CSF and PBMCs. Day 14 PBMC data is not shown due to fitting a full model for column effect, row effect, and column/row interaction effect. *P<0.05; **P<0.01; ns = not significant. n = 6 patients with matched CSF and PBMC samples. Total samples: CSF, n = 23; PBMC, n = 19

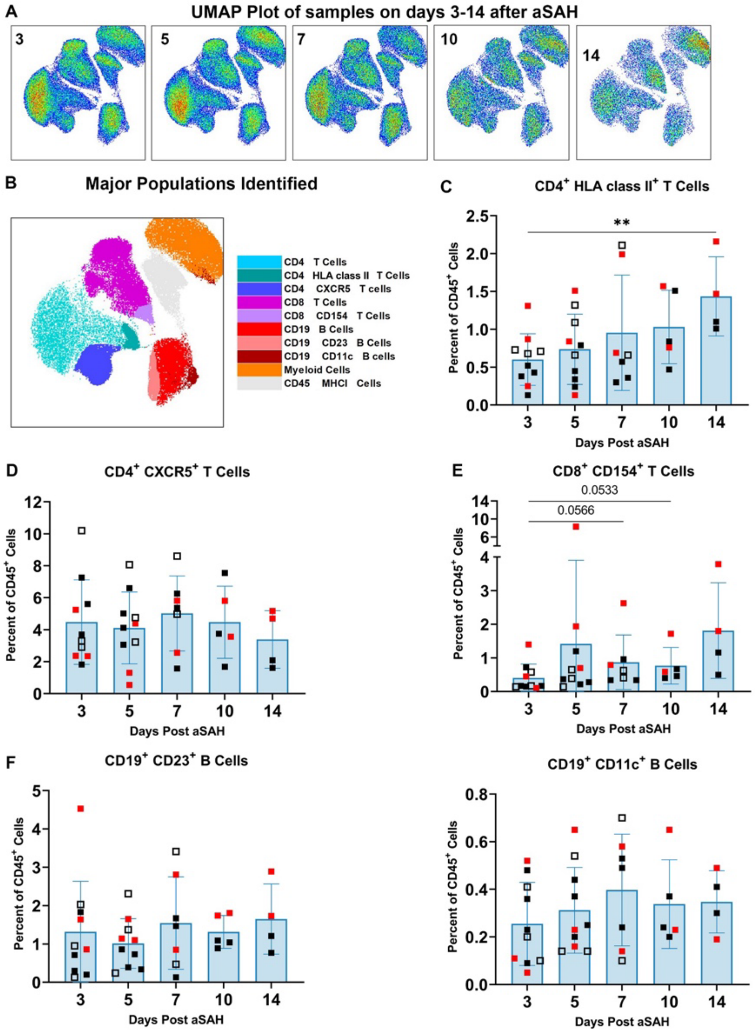


**Supplemental Figure 3: Specific T and B cell subpopulations from aSAH patient PBMC samples over 14 days.** A) Flow cytometry UMAP plot of all samples across a 14-day time span. B) Major populations identified. C) CD4+ MHCII+ T cells. D) CD4+ CXCR5+ T cells. E) CD8+ CD154+ T cells. F) CD19+ CD23+ B cells and CD19+ CD11c+ B cells. Bar graphs represent mean values; error bars indicate SD. Cell populations are shown as a percentage of live CD45+ cells. Hollow squares indicate patients without vasospasm, filled squares indicate patients with vasospasm, and red squares indicate patients with vasospasm and delayed cerebral ischemia (DCI). Statistical analysis: A one-way ANOVA (mixed-effects model REML) with Geisser-Greenhouse correction was performed, followed by Dunnett's multiple comparisons test comparing the means of each day to the mean from day 3. *P<0.05; **P<0.01. Data from 10 patients, n = 36 samples.

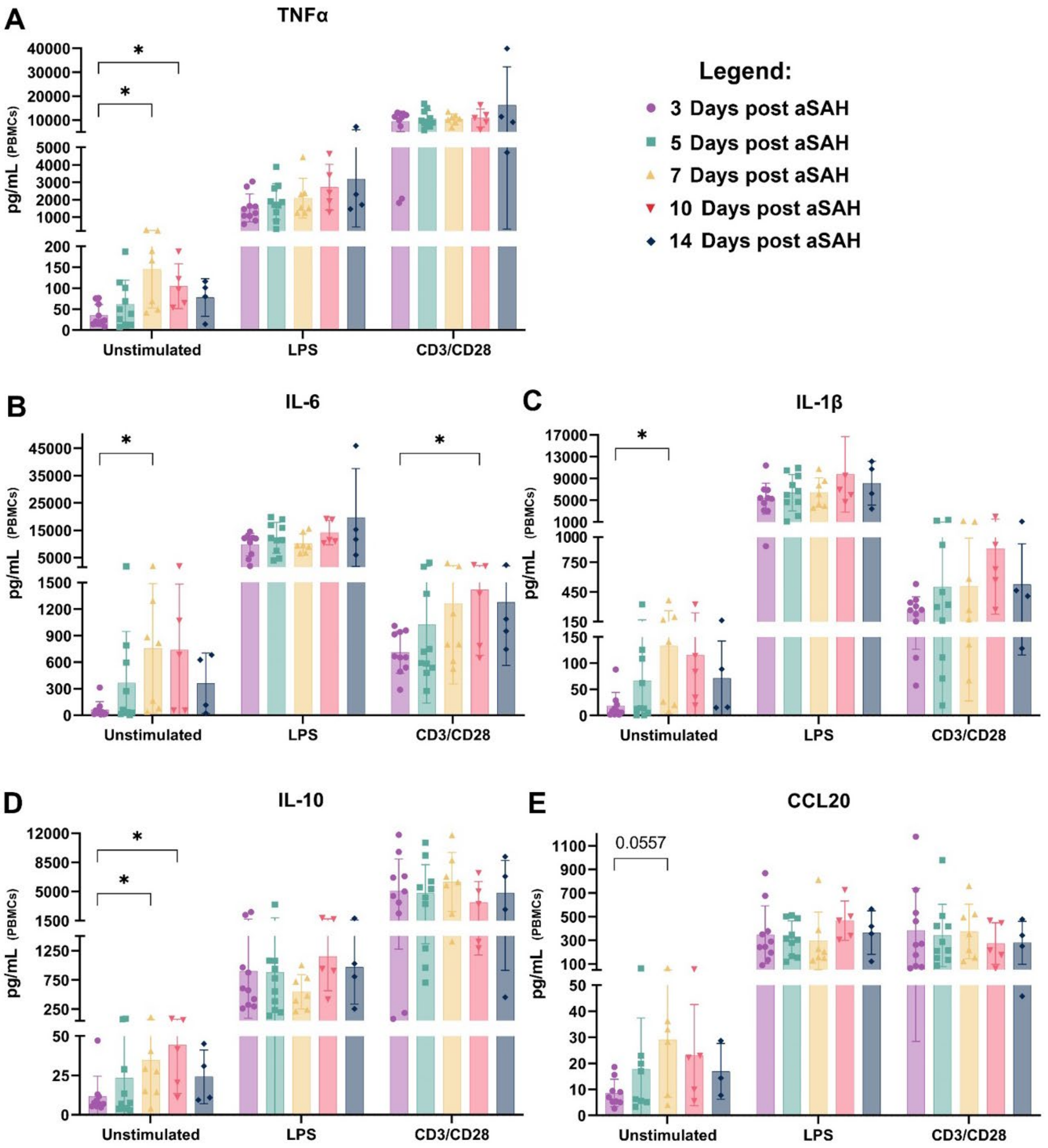


**Supplemental Figure 4: Select PBMC cytokines over days 3-14 comparing cytokine production in unstimulated, LPS, and CD3/CD28 stimulated conditions.** A) TNFα B) IL-6 C) IL-1β D) IL-10 E) CCL20. We conducted a 2-way ANOVA or mixed-effects model (REML). We tested fixed effects (type III) for stimulation, days post-aSAH, and Stimulation x days post-aSAH. Stimulation was significant to at least P<0.01 in all cases, while days post-aSAH, and Stimulation x days post-aSAH did not yield significance. Additionally, we performed Fisher's LSD for a planned comparison of cytokine levels at different days against the first collection time point. *P<0.05; **P<0.01. Data shown is from 10 patients, with n = 108 samples or n = 36 per condition (Unstimulated, LPS, CD3/CD28).

**Supplemental Table 1: Flow Cytometry Key Resource Table, Extracellular Panel**

| Antibody | Fluorophore | Isotype/Clone | Vendor | Catalog number |
|---|---|---|---|---|
| Fixable Viability Dye 780 | APC-Cy7 | N/A | BD | 565388 |
| CD45 | BUV805 | HI30 | BD Horizon | 612891 |
| CD19 | PE-Cy7 | HIB19 | TONBO | 60-0199-T100 |
| CD3 | BV480 | UCHT1 | BD Horizon | 566105 |
| CD4 | BUV396 | M-T477 | BD Biosciences | 742738 |
| CD8 | BV650 | RPA-T8 | BD Biosciences | 563821 |
| CD11b | BB515 | ICRF44 | BD Horizon | 564517 |
| CD11c | BUV661 | B-ly6 | BD Horizon | 612967 |
| CD14 | BUV737 | M5E2 | BD Horizon | 612763 |
| CD66b | BV421 | G10F5 | BD Horizon | 562940 |
| CD138 | BV711 | MI15 | BD Horizon | 583184 |
| CXCR3 (CD183) | BUV496 | 1C6/CXCR3 | BD Biosciences | 741178 |
| **CD161** | **PE** | **HP-3G10** | **BD Biosciences** | **566843** |

**Supplemental Table 2: Flow Cytometry Key Resource Table, Extracellular Panel II**

| Antibody | Fluorophore | Isotype/Clone | Vendor | Catalog number |
|---|---|---|---|---|
| Fixable Viability Dye 780 | APC-Cy7 | N/A | BD | 565388 |
| CD45 | BUV805 | HI30 | BD Horizon | 612891 |
| CD19 | BUV563 | SJ25C1 | BD | 612916 |
| CD3 | BV480 | UCHT1 | BD Horizon | 566105 |
| CD4 | BUV396 | SK3 | BD | 563550 |
| CD8b | BV650 | RPA-T8 | BD | 742393 |
| CD11b | BV711 | ICRF44 | BD | 740771 |
| CD11c | PE-Cy7 | B-ly6 | BD | 561356 |
| CD25 | BV421 | BC96 | BD | 567485 |
| CD23 | BV605 | M-L233 | BD | 740414 |
| CCR7 | BUV496 | 2-LI-A | BD | 749827 |
| CXCR5 | R718 | RF8B2 | BD | 752012 |
| CCR3 (CD193) | APC | 5 E8 | BD | 558208 |
| CD154 (CD40L) | PE-CF594 | TRAP1 | BD Horizon | 563589 |
| IL-21R | BB700 | 2SX21R | Invitrogen | 46-3601-42 |
| CD69 | BV786 | FH50 | BD Horizon | 563834 |
| MHC I (HLA I) | BB515 | W6/32 | Biolegend | 311404 |
| **MHC II (HLA DR DQ DP)** | **PE** | **TU39** | **Biolegend** | **361716** |
